# Single-Dose Suprachoroidal AXT107 in Neovascular Age-Related Macular Degeneration: The Phase 1/2a DISCOVER Trial

**DOI:** 10.64898/2026.09.19.26363031

**Authors:** Niranjan B. Pandey, Adam C. Mirando, Thomas J. Robinson, David R. P. Almeida, Sabin Dang, David R. Lally, William Z. Bridges, Peter A. Campochiaro, Aleksander S. Popel, Jordan J. Green

## Abstract

Frequent injections remain a substantial burden in the treatment of neovascular age-related macular degeneration (nAMD). AXT107 (gersizangitide) is a long-acting, integrin-disrupting peptide designed for suprachoroidal administration. In the phase 1/2a, open-label DISCOVER trial, 15 participants with active nAMD received a single suprachoroidal injection of AXT107 microparticles at 0.125, 0.25, or 0.5 mg and were followed for 40 weeks. Safety and tolerability were the primary outcomes; visual acuity, retinal thickness, and time to protocol-specified rescue anti-vascular endothelial growth factor treatment were exploratory outcomes. No AXT107-related serious adverse events, clinically significant intraocular inflammation, or sustained intraocular pressure elevations were observed. Fourteen eyes met the protocol-specified baseline imaging criterion for efficacy analyses. Among these eyes, two treatment-experienced eyes completed 40 weeks without protocol-specified rescue, and a third first received protocol-specified rescue at week 36. Anatomical improvements were observed in at least one eye in each dose cohort, although responses varied. A single suprachoroidal injection of AXT107 was well tolerated in this small first-in-human study. The exploratory findings warrant evaluation in a controlled trial

**One Sentence Summary:** Suprachoroidal AXT107 was well tolerated, with some eyes remaining free of protocol-specified rescue for up to 40 weeks.

## Introduction

Neovascular age-related macular degeneration (nAMD) is a prevalent, progressive cause of central vision loss and blindness in older adults (1). Although it accounts for a minority of AMD cases, the neovascular form is responsible for most AMD-related severe visual impairment because of its rapid onset and exudative course (1). Intravitreal anti-vascular endothelial growth factor (VEGF) therapy has transformed the prognosis and treatment of nAMD, with randomized trials demonstrating substantial gains or stabilization of vision when strict monthly or treat-and-extend regimens are followed (2–5). Population-level data have also shown that widespread use of anti-VEGF agents results in reductions in nAMD-related blindness (6). In routine practice, however, injection intervals are often longer, visit adherence is lower, and cumulative treatment exposure is substantially less than in registration trials (5), resulting in treatment gaps. These gaps in treatment have been associated with smaller gains in visual acuity, loss of earlier improvements over time, and incomplete symptom control (5). Together, these observations highlight an unmet medical need for therapies that are efficacious under real-life conditions and provide durable disease control with fewer injections and work by mechanisms that go beyond VEGF neutralization (5,7).

Currently approved therapies for nAMD, including ranibizumab, aflibercept, brolucizumab, and the bispecific antibody faricimab, target VEGFA alone or VEGFA together with angiopoietin□2 (Ang2), and have reduced nAMD□related blindness (2,3,8). However, these agents typically require frequent intravitreal (IVT) injections, often every 4-8 weeks, to maintain disease control, imposing a substantial treatment burden on patients, caregivers, and health systems with worse outcomes in the real world compared with results from clinical trials (5,7). Moreover, many patients show persistent or recurrent exudation despite intensive anti VEGF therapy, and long term treatment has been associated in several studies with increased macular/geographic atrophy, although causality remains debated, highlighting the need for therapies that require less frequent injections and address mechanisms that go beyond VEGF-dependent ones (5,7,9,10).

The angiopoietin Tie2 pathway is increasingly regarded as a critical regulator of vascular stability, permeability, and inflammation in the retina and choroid (11,12). In health, Ang1 mediated activation of the endothelial receptor tyrosine kinase Tie2 promotes junctional integrity, pericyte support, and barrier function, whereas in disease states such as nAMD and diabetic macular edema, high levels of Ang2 antagonize Tie2, destabilize vessels, and enhance leakage and inflammatory signaling (11–13). Dual inhibition of VEGFA and Ang2 with faricimab has validated Ang2 as a therapeutic target, improving anatomical outcomes and permitting extended dosing in some patients compared with VEGFA inhibition alone (8,14,15). Notably, Ang2 is not simply a passive antagonist of Tie2 but can act as a context-dependent partial or full agonist under certain conditions, such as when Tie2 is appropriately clustered or presented at the cell surface, raising the possibility that the same disease-upregulated Ang2 pool driving vascular destabilization could instead be redirected to activate rather than antagonize Tie2 (16,17). Nevertheless, faricimab acts primarily by neutralizing Ang2, and its mechanism does not allow for Ang2 itself to serve as an abundant, disease upregulated ligand to drive Tie2 activation and vascular stabilization (11,12,15).

AXT107 (Gersizangitide) is a 20□amino□acid peptide derived from type IV collagen that binds integrins α5β1 and αvβ3 and acts through multiple mechanisms on the ocular vasculature (18,19). In preclinical studies, AXT107 disrupted integrin-receptor complexes, inhibited VEGFR2 and other pro□angiogenic receptor tyrosine kinases, reduced neovascularization, and markedly suppressed VEGF□induced vascular leakage (18–20). In parallel, AXT107 induced re-localization and clustering of Tie2 at endothelial cell-cell junctions and converted Ang2 from an antagonist into a potent agonist of Tie2, resulting in high levels of Tie2 phosphorylation, Akt activation, reorganization of cortical actin, and tightening of endothelial junctions *in vitro* and *in vivo* (20,21). In models of ocular inflammation, AXT107 used endogenously released Ang2 to activate Tie2 and inhibit inflammation by reducing expression of VCAM□1 and ICAM□1, leukostasis, and vascular leakage (21). Together, these data support that AXT107 can simultaneously inhibit VEGF signaling, activate Tie2 using elevated Ang2, and suppress inflammation, offering a more comprehensive approach and different mechanism to treating nAMD than VEGFA or VEGFA/Ang2 neutralization alone (18,20,21).

An additional advantageous property of AXT107 is its intrinsic ability to self□assemble without the need for an exogenous biomaterial carrier (22). The peptide is soluble at low pH and low ionic strength, but upon exposure to the physiological pH and ionic strength of the vitreous it precipitates and, because of the high viscosity of the vitreous, coalesces in place into a compact gel like mass that remains below the visual axis and slowly dissolves over months, with an apparent intravitreal half□life of approximately 180 days in animal studies (data on file, Johns Hopkins University School of Medicine from AsclepiX Therapeutics). In rabbit models, a single intraocular injection of AXT107 produced prolonged suppression of VEGF induced vascular leakage and neovascularization, with durability exceeding that of aflibercept (18). The same gel forming property was leveraged to create a microparticle formulation suitable for suprachoroidal delivery: controlled mixing of a low pH, low ionic strength AXT107 solution with a higher□pH, higher□ionic□strength solution yields self□assembled peptide microparticles in which the drug itself forms the particulate depot, without encapsulating polymers or other structural excipients (22). The multifactorial and long-acting characteristics of AXT107 support its evaluation as a potential therapy designed to extend treatment intervals while addressing barrier dysfunction and inflammation in addition to VEGF-driven angiogenesis.

Early clinical development of AXT107 used an intravitreal self□forming gel formulation, initially evaluated in a Phase 1 diabetic macular edema trial (NCT04697758) and, after acceptable safety in the initial low dose cohort, in a Phase 1 nAMD trial (NCT04746963) using the same intravitreal formulation (23,24). Dose□related sustained elevations in intraocular pressure (IOP) that emerged only after dose escalation in the DME trial and initiation of the nAMD trial, and were not adequately controlled with pressure lowering medications, raised concern that intravitreal microscopic gel fragments might obstruct aqueous outflow at the trabecular meshwork. These observations motivated reformulation of AXT107 as a microparticle suspension for suprachoroidal administration. This approach was intended to deliver drug directly to the choroid and outer retina while minimizing exposure to the vitreous and anterior chamber, thereby reducing the risk of outflow obstruction and intraocular pressure elevation. In preclinical studies, suprachoroidal administration of AXT107 microparticles inhibited new blood vessel growth in a rat choroidal neovascularization model (22), and a single dose GLP toxicology study in minipigs demonstrated acceptable ocular safety at doses up to 1.25 mg (22).

Based on this mechanistic rationale and translational safety and efficacy data, we conducted DISCOVER (NCT05859776), a Phase 1/2a, dose-escalation trial to evaluate the safety, tolerability, and preliminary bioactivity of a single suprachoroidal injection of AXT107 microparticles in eyes with nAMD. The study employed stepwise dose escalation (0.125, 0.25, and 0.5 mg), starting at one-tenth of the highest dose shown to be safe in minipigs, followed by expansion of the highest dose cohort, including treatment-naïve eyes with newly diagnosed nAMD. Each subject received only one suprachoroidal injection and was followed for 40 weeks, with protocol-specified intravitreal anti-VEGF rescue allowed from week 12. Investigator-initiated anti-VEGF treatment could occur before Week 12 when clinically indicated. In this report, we describe the safety profile, single-dose durability, and exploratory efficacy outcomes from DISCOVER, representing the first-in-human evaluation of a multimodal (VEGFA pathway and angiopoietin Tie2 pathway) long-acting therapy for nAMD delivered to the suprachoroidal space.

## Results

### Subject enrollment and baseline characteristics

Between January and May 2024, 20 subjects were screened at 4 U.S. clinical sites, 5 were screen failures, and 15 subjects with nAMD were enrolled and received a single suprachoroidal injection of AXT107 (**Fig. 1**). Three subjects received 0.125 mg (Cohort 1), 3 received 0.25 mg (Cohort 2), and 9 received 0.5 mg (Cohort 3), including 6 additional subjects in the cohort 3 expansion (**Table 1**). The enrolled population had a mean age of 80 years (range, 72-90), included 8 females and 7 males, and consisted entirely of White, non-Hispanic subjects. Among the 14 eligible eyes included in efficacy analyses, baseline mean CST was 504.4 µm and mean BCVA was 54.1 ETDRS letters. Among the 9 eyes treated with 0.5 mg, 5 were treatment-experienced partial responders and 4 were treatment-naïve eyes with newly diagnosed nAMD. Fourteen of 15 subjects completed the full 40-week study; 1 mid-dose subject exited at week 32 after relocation for non-safety reasons.

**Fig. 1.**
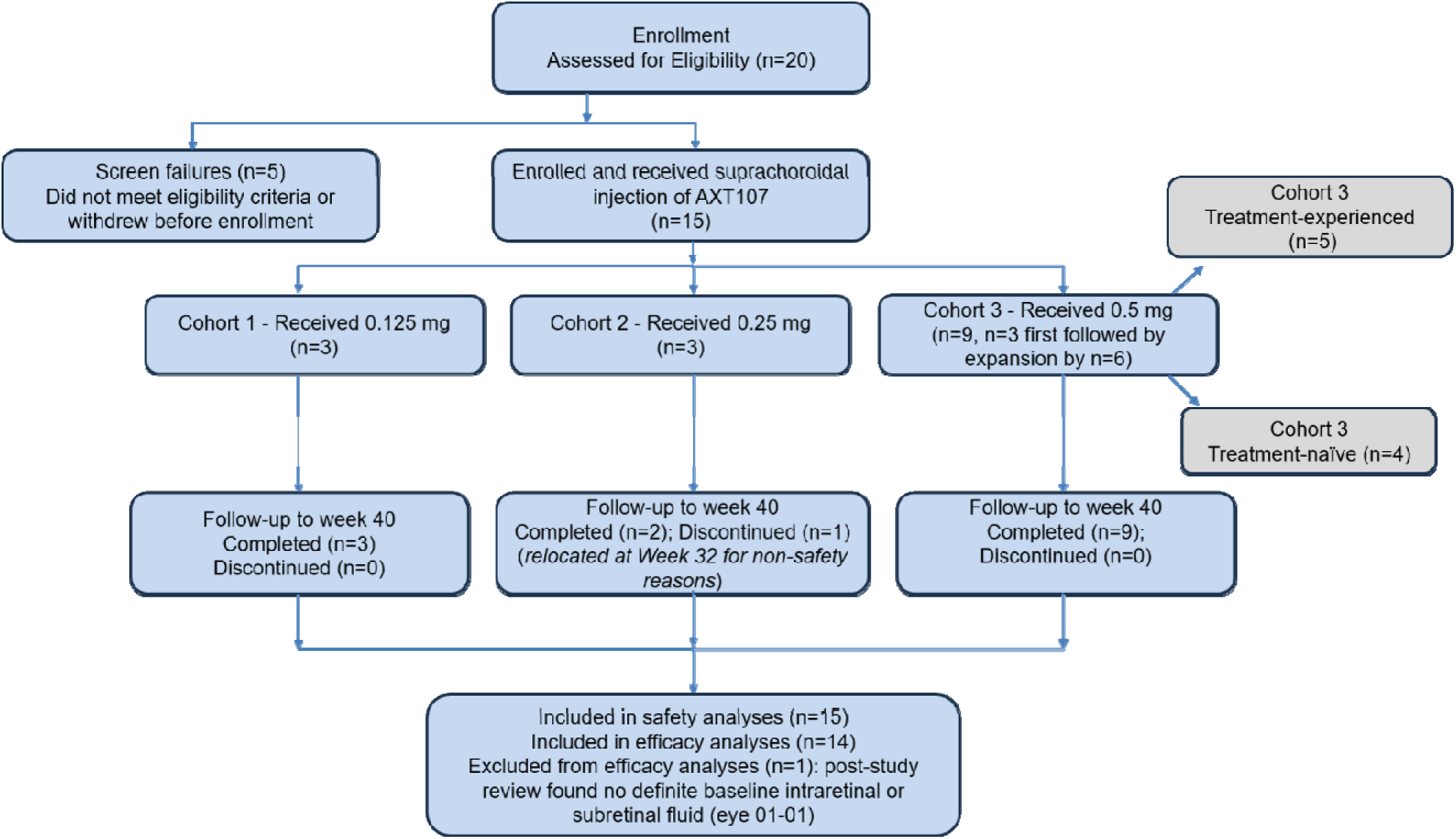
Participant disposition, dose cohorts, and analysis populations in the Phase 1/2a DISCOVER trial. Of 20 subjects assessed for eligibility, 15 received a single suprachoroidal injection of AXT107: 3 received 0.125 mg, 3 received 0.25 mg, and 9 received 0.5 mg. All 15 treated eyes were included in safety analyses. Following post-study review of screening and baseline SD-OCT images, treatment-experienced Eye 03 did not meet the protocol-specified imaging eligibility criterion and was excluded from efficacy analyses; 14 protocol-eligible treated eyes were included in efficacy analyses. One subject in the 0.25-mg cohort discontinued at Week 32 because of relocation for non-safety reasons; all other subjects completed follow-up through Week 40.

**Table 1.** Baseline demographic and ocular characteristics of the study eye.

| Cohort | De-identified Eye Label | Age Range | Previous Anti-VEGF Injections*, n | Pre-treatment Status | Lens Status | Duration of nAMD | CNV Location | CNV Type | BCVA** (letters) | CST (µm) |
| --- | --- | --- | --- | --- | --- | --- | --- | --- | --- | --- |
| 1 | Eye 01 | 75-79 | 6 | Treatment-experienced | Phakic | 3 yr | Subfoveal | Minimally Classic/Occult | 51 | 447 |
|  | Eye 02 | 75-79 | 2 | Treatment-experienced | Phakic | 3 yr | Subfoveal | Predominantly Classic | 46 | 474 |
|  | Eye 03 | 80-84 | 4 | Treatment-experienced | Pseudophakic | 10 yr | Subfoveal | Minimally Classic/Occult | 65 | 312 |
| 2 | Eye 04 | 80-84 | 4 | Treatment-experienced | Pseudophakic | 5 yr | Subfoveal | Predominantly Classic | 32 | 927 |
|  | Eye 05 | 85-89 | 2 | Treatment-experienced | Phakic | 1 yr | Subfoveal | Predominantly Classic | 44 | 352 |
|  | Eye 06 | 80-84 | 5 | Treatment-experienced | Phakic | 5 yr | Juxtafoveal | Minimally Classic/Occult | 52 | 384 |
| 3<br>Treatment-experienced | Eye 07 | 85-89 | 2 | Treatment-experienced | Pseudophakic | 2 yr | Subfoveal | Minimally Classic/Occult | 65 | 780 |
|  | Eye 08 | 90-94 | 2 | Treatment-experienced | Pseudophakic | 2 yr | Subfoveal | Predominantly Classic | 60 | 446 |
|  | Eye 09 | 70-74 | 3 | Treatment-experienced | Phakic | 3 yr | Subfoveal | Minimally Classic/Occult | 65 | 400 |
|  | Eye 10 | 80-84 | 4 | Treatment-experienced | Pseudophakic | 4 yr | Subfoveal | Minimally Classic/Occult | 56 | 414 |
|  | Eye 11 | 75-79 | 3 | Treatment-experienced | Phakic | < 1 yr | Subfoveal | Minimally Classic/Occult | 54 | 358 |
| 3<br>Treatment-Naïve | Eye 12 | 80-84 | 0 | Treatment-naïve | Pseudophakic | Newly diagnosed | Subfoveal | Minimally Classic/Occult | 47 | 455 |
|  | Eye 13 | 75-79 | 0 | Treatment-naïve | Pseudophakic | Newly diagnosed | Subfoveal | Minimally Classic/Occult | 69 | 637 |
|  | Eye 14 | 70-74 | 0 | Treatment-naïve | Pseudophakic | Newly diagnosed | Subfoveal | Predominantly Classic | 52 | 600 |
|  | Eye 15 | 70-74 | 0 | Treatment-naïve | Phakic | Newly diagnosed | Juxtafoveal | Predominantly Classic | 64 | 388 |

\*The number of anti-VEGF injections during the 12 months before baseline is shown.
\*\* BCVA values shown are from the baseline visit. Treatment-naïve Eye 13 had a screening BCVA of 65 ETDRS letters, meeting the protocol-specified eligibility criterion; the baseline BCVA was 69 ETDRS letters approximately 2 weeks later.

### Safety and tolerability

No serious adverse events related to AXT107 occurred, and no cases of endophthalmitis, retinal detachment, suprachoroidal hemorrhage, or clinically significant intraocular inflammation were observed through week 40. Ocular treatment emergent adverse events (TEAEs) were generally mild or moderate, with no subjects discontinued due to ocular TEAEs and no serious ocular TEAEs (**Table 2A**). Non□ocular TEAEs were infrequent, predominantly mild, and not considered related to AXT107; no deaths, life□threatening events, or hospitalizations occurred, and no subjects were withdrawn because of non□ocular TEAEs (**Table 2B**). No clinically meaningful intraocular pressure (IOP) elevations occurred at any dose, and no subject required chronic IOP lowering therapy.

**Table 2.** Safety findings. A. Overview of ocular treatment-emergent adverse events (TEAEs) in the study eye through Week 40 B. Overview of non□ocular TEAEs through Week 40

| Parameter | 0.125 mg<br>(N=3) | 0.25 mg<br>(N=3) | 0.5 mg<br>(N=9) | Overall<br>(N=15) |
| --- | --- | --- | --- | --- |
| Subjects with any ocular TEAE, n (%) | 2 (66.7%) | 1 (33.3%) | 5 (55.6%) | 8 (53.3%) |
| Total number of ocular TEAEs, n | 4 | 1 | 9 | 14 |
| Subjects with ocular serious TEAEs, n (%) | 0 (0%) | 0 (0%) | 0 (0%) | 0 (0%) |
| Subjects withdrawn due to ocular TEAEs, n (%) | 0 (0%) | 0 (0%) | 0 (0%) | 0 (0%) |
| Subjects with ocular TEAEs by maximum severity |  |  |  |  |
| – Mild, n (%) | 1 (33.3%) | 1 (33.3%) | 3 (33.3%) | 5 (33.3%) |
| – Moderate, n (%) | 1 (33.3%) | 0 (0%) | 2 (22.2%) | 3 (20.0%) |
| – Severe, n (%) | 0 (0%) | 0 (0%) | 0 (0%) | 0 (0%) |
| Subjects with ocular TEAEs by maximum relationship* to AXT107 |  |  |  |  |
| – Unrelated, n (%) | 2 (66.67%) | 1 (33.3%) | 4 (44.4%) | 7 (46.67%) |
| – Related, n (%) | 0 (0%) | 0 (0%) | 1 (11.1%) | 1 (6.7%) |

\* Relationship is the investigator's assessment of causality (unrelated = not or unlikely related; related = possibly, probably, or definitely related). Each subject is counted once according to their maximum (most related) TEAE.

B. Overview of non-ocular TEAEs through Week 40
| Parameter | 0.125 mg<br>(N=3) | 0.25 mg<br>(N=3) | 0.5 mg<br>(N=9) | Overall<br>(N=15) |
| --- | --- | --- | --- | --- |
| Subjects with any non-ocular TEAE, n (%) | 1 (33.3%) | 0 (0%) | 4 (44.4%) | 5 (33.3%) |
| Total number of non-ocular TEAEs, n | 1 | 0 | 8 | 9 |
| Subjects with non-ocular serious TEAEs, n (%) | 0 (0%) | 0 (0%) | 0 (0%) | 0 (0%) |
| Death/life-threatening/hospitalization | 0 (0%) | 0 (0%) | 0 (0%) | 0 (0%) |
| Subjects withdrawn due to non-ocular TEAEs, n (%) | 0 (0%) | 0 (0%) | 0 (0%) | 0 (0%) |
| Subjects with non-ocular TEAEs by maximum severity |  |  |  |  |
| - Mild, n (%) | 1 (33.3%) | 0 (0%) | 2 (22.2%) | 3 (20.0%) |
| - Moderate, n(%) | 0 (0%) | 0 (0%) | 2 (22.2%) | 2 (13.3%) |
| - Severe, n(%) | 0 (0%) | 0 (0%) | 0 (0%) | 0 (0%) |
| Subjects with non-ocular TEAEs by maximum relationship* to AXT107 |  |  |  |  |
| - Unrelated, n (%) | 1 (33.3%) | 0 (0%) | 4 (44.4%) | 5 (33.3%) |
| - Related, n (%) | 0 (0%) | 0 (0%) | 0 (0%) | 0 (0%) |

\* Relationship is the investigator's assessment of causality (unrelated = not or unlikely related; related = possibly, probably, or definitely related). Each subject is counted once according to their maximum (most related) TEAE.

### BCVA outcomes

BCVA at each study visit is shown for the 14 eligible eyes included in efficacy analyses in **Fig. 2A**, with complete datasets and datasets censored after first anti-VEGF treatment displayed in parallel. Because anti-VEGF treatment, including investigator-initiated treatment before Week 12 and protocol-specified rescue from Week 12, could occur during follow-up, interpretation of late BCVA values requires distinction between the AXT107 treatment period and periods after anti-VEGF treatment. For descriptive analyses, the AXT107 treatment period was defined as the interval from AXT107 administration through the assessment obtained immediately before the first anti-VEGF injection, whether investigator-initiated or administered as protocol-specified rescue. Several eyes maintained visual acuity for prolonged periods during AXT107 treatment, whereas in other eyes later BCVA improvement occurred only after subsequent anti-VEGF treatment.

**Fig. 2.**
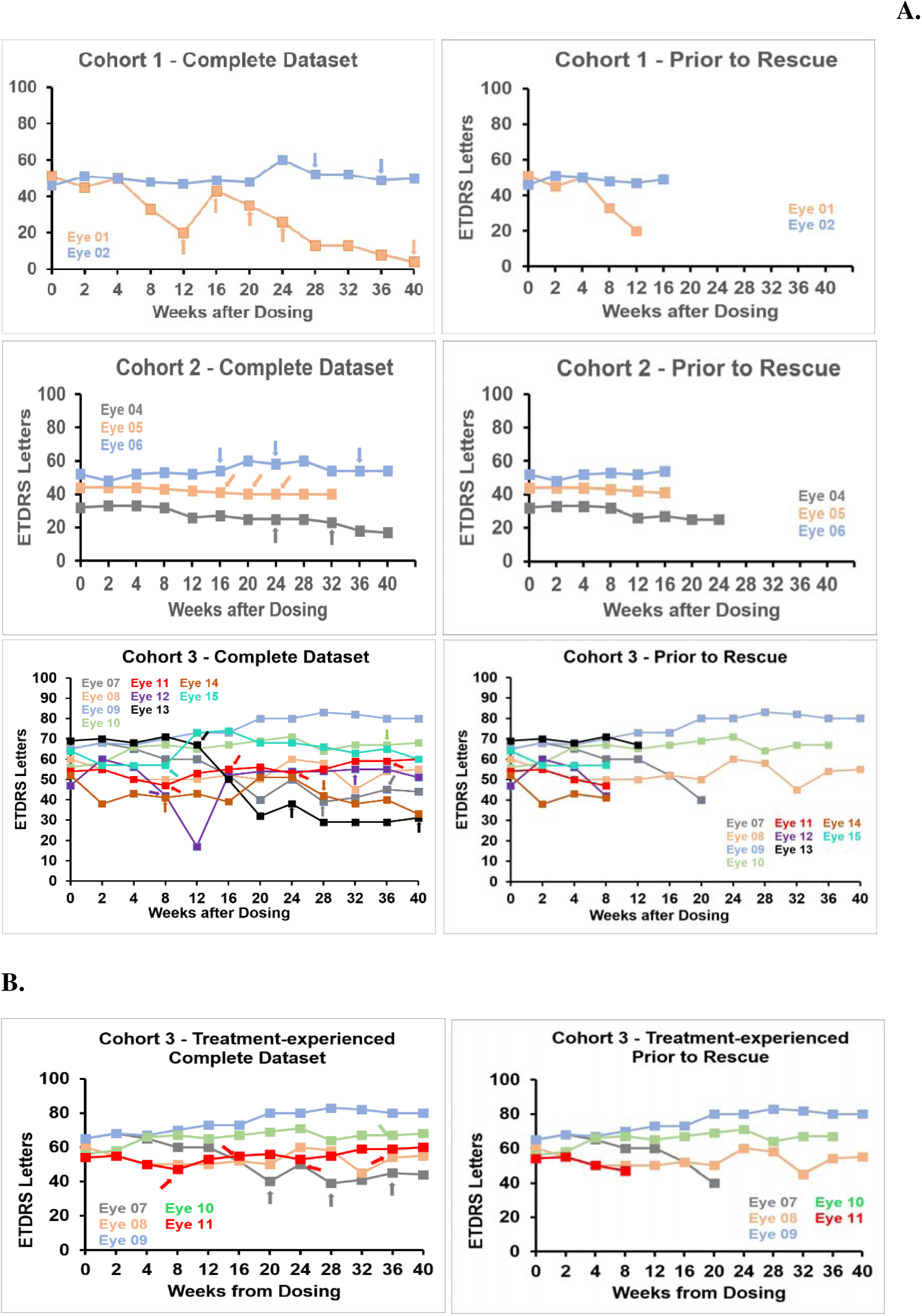
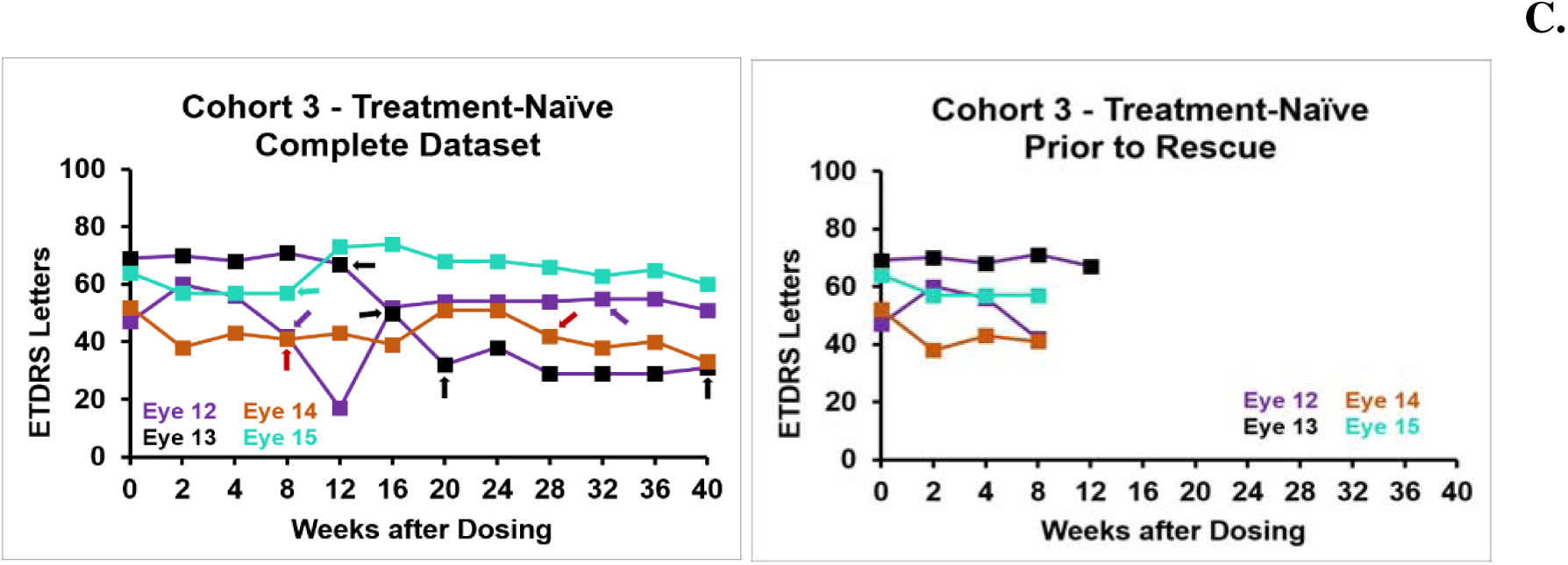
BCVA outcomes after a single suprachoroidal injection of AXT107. (A) BCVA in the 14 eligible eyes by cohort and dose. (B) BCVA in treatment-experienced eyes in Cohort 3 (0.5 mg). (C) BCVA in treatment-naïve eyes in Cohort 3 (0.5 mg). In each of A, B, and C, the left panel shows the complete dataset, including visits after the first anti-VEGF treatment, and the right panel shows values through the first anti-VEGF treatment, with subsequent visits censored. Arrows indicate anti-VEGF injections; treatment before Week 12 was investigator initiated, whereas protocol-specified rescue was permitted from Week 12 onward according to prespecified BCVA or CST criteria.

Among treatment-experienced eyes in the 0.5-mg Cohort 3, Eyes 08 and 09 had stable or improved BCVA with no anti-VEGF treatment through Week 40, and Eye 10 had stable vision without anti-VEGF treatment through Week 36 (**Fig. 2B**). In contrast, treatment-experienced Eye 11 received investigator-initiated anti-VEGF treatment at Week 8, followed by anti-VEGF injections at Weeks 16, 24, and 36; accordingly, late BCVA improvement in this eye cannot be attributed to AXT107 alone. Treatment-experienced Eye 07 likewise required rescue at Weeks 20, 28, and 36, so its later trajectory reflects combined exposure to AXT107 and rescue therapy rather than AXT107 alone.

In treatment-naïve 0.5-mg eyes (**Fig. 2C**), BCVA responses were more variable and rescue generally occurred earlier. Eye 12 received investigator-initiated anti-VEGF treatment at Week 8 and protocol-specified rescue at Week 32; Eye 14 received investigator-initiated treatment at Week 8 and protocol-specified rescue at Week 28; and Eye 15 received investigator-initiated treatment at Week 8. Thus, for most treatment-naïve eyes, only the early portion of the BCVA curve reflects treatment with AXT107. In exploratory supplemental analysis, mean BCVA change was more favorable while eyes remained on AXT107 than during subsequent periods on rescue anti-VEGF therapy (**Fig. S1**), although this comparison is descriptive and affected by the fact that rescue was given to worsening eyes.

### Exploratory BCVA responder analysis

At the last on□treatment visit (week 40 for eyes without rescue and the rescue□day visit for eyes with rescue), BCVA changes from baseline ranged from +15 to –31 ETDRS letters among the 14 eligible eyes included in efficacy analyses. In this exploratory analysis, 3 eyes gained ≥5 letters, 3 gained ≥10 letters, and 1 gained ≥15 letters at this endpoint, reflecting modest responder proportions consistent with the small, heterogeneous phase 1/2a population. These descriptive findings parallel the subject-level trajectories: several eyes maintained BCVA during the AXT107-treatment period, whereas some eyes that received subsequent anti-VEGF treatment had larger BCVA losses by the end of follow-up (**Fig. S1**).

### CST outcomes

CST over time is shown for the 14 eligible eyes included in efficacy analyses in **Fig. 3A** and for treatment-experienced and treatment-naïve eyes in the 0.5-mg cohort in **Fig. 3B** and **Fig. 3C**, respectively. Anatomical improvement was observed in at least 1 eye in each dose cohort. Treatment-experienced Eyes 01 and 02 first received protocol-specified rescue at Weeks 12 and 16, respectively; all Cohort 2 eyes received anti-VEGF treatment between Weeks 16 and 24. Subsequent anti-VEGF injections were administered at Weeks 20 and 24 for treatment-experienced Eye 05, at Weeks 24 and 36 for treatment-experienced Eye 06, and at Week 32 for treatment-experienced Eye 04.

**Fig. 3.**
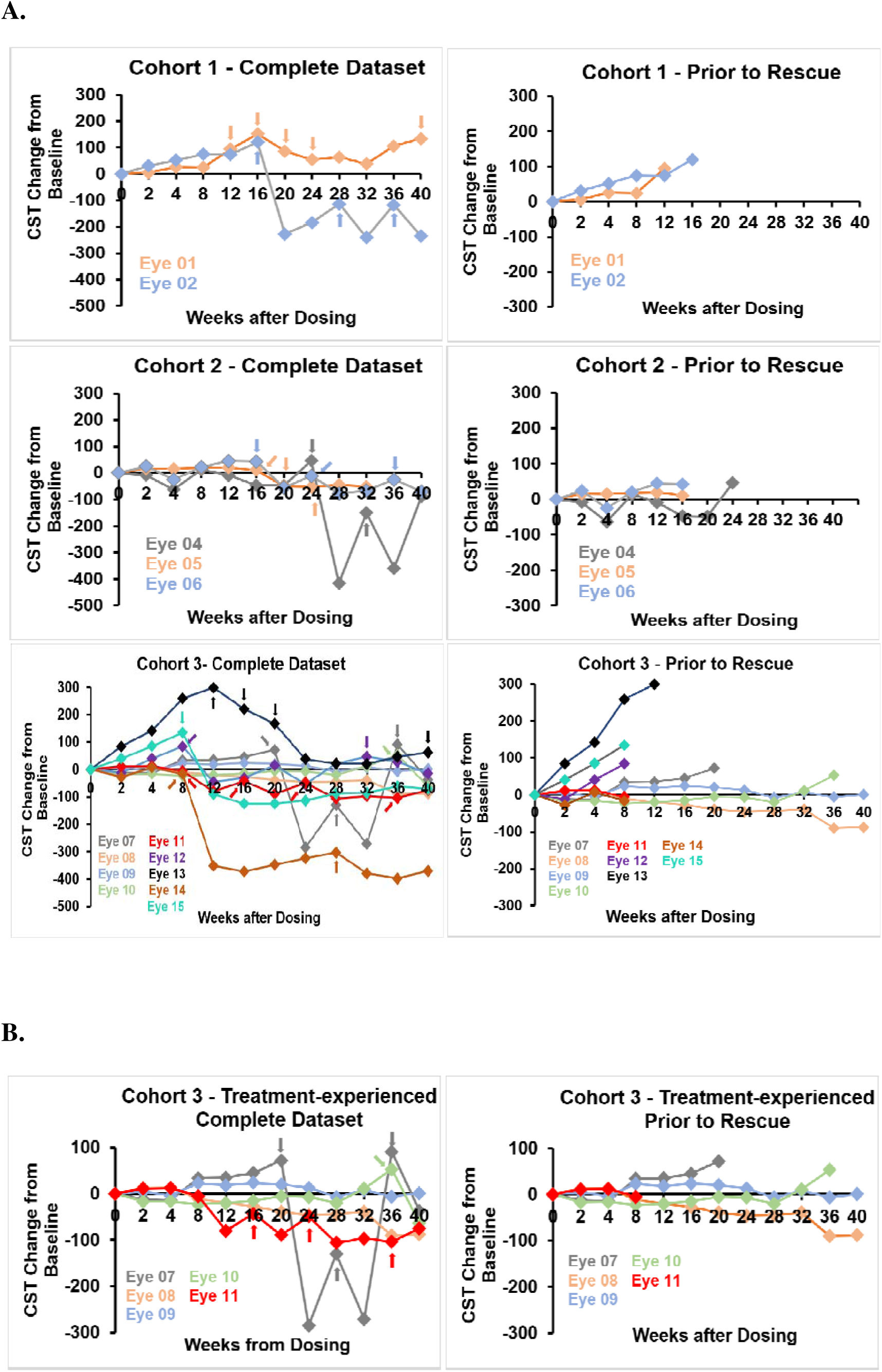
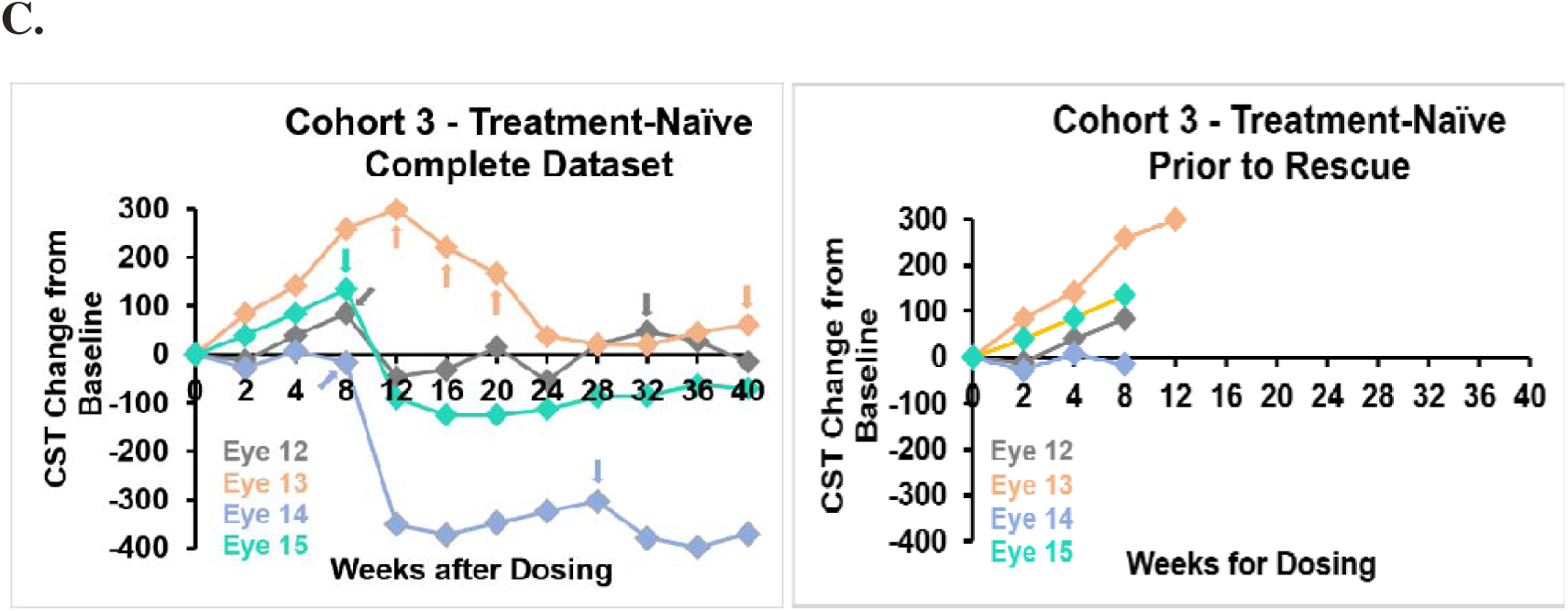
CST changes after a single suprachoroidal injection of AXT107. (A) Individual change from baseline in CST in the 14 eligible eyes by cohort and dose. (B) CST changes in treatment-experienced eyes in Cohort 3 (0.5 mg). (C) CST changes in treatment-naïve eyes in Cohort 3 (0.5 mg). In each of A, B, and C, the left panel shows the complete dataset, including visits after the first anti-VEGF treatment, and the right panel shows values through the first anti-VEGF treatment, with subsequent visits censored. Arrows indicate anti-VEGF injections; treatment before Week 12 was investigator initiated, whereas protocol-specified rescue was permitted from Week 12 onward according to prespecified BCVA or CST criteria.

In the 0.5-mg cohort, treatment-experienced eyes generally showed the greatest CST stability: Eyes 08 and 09 had stable or improved CST at Week 40 with no anti-VEGF treatment, and Eye 10 showed a sustained reduction in CST relative to baseline through Week 28, followed by a gradual increase that preceded rescue at Week 36. Treatment-experienced Eye 11 and three treatment-naïve eyes received investigator-initiated anti-VEGF treatment at Week 8; the remaining treatment-naïve eye first received protocol-specified rescue at Week 12. The curves censored after first anti-VEGF treatment in **Fig. 3B** and **Fig. 3C** help distinguish CST changes during AXT107 treatment from those occurring after anti VEGF treatment.

### Durability and rescue

**Fig. 4** shows the time to first anti-VEGF treatment or censoring among the 14 eligible eyes included in efficacy analyses. Two eligible treatment-experienced eyes, Eyes 08 and 09, required no protocol-specified rescue anti-VEGF through Week 40, and treatment-experienced Eye 10 first required protocol-specified rescue at Week 36. Most other eyes received first anti-VEGF treatment between Weeks 8 and 24; treatment at Week 8 was investigator initiated, whereas first treatment from Week 12 onward was protocol-specified rescue, earlier first anti-VEGF treatment generally occurring in treatment naïve eyes and later rescue more common in treatment-experienced eyes in the 0.5 mg cohort.

**Fig. 4.**
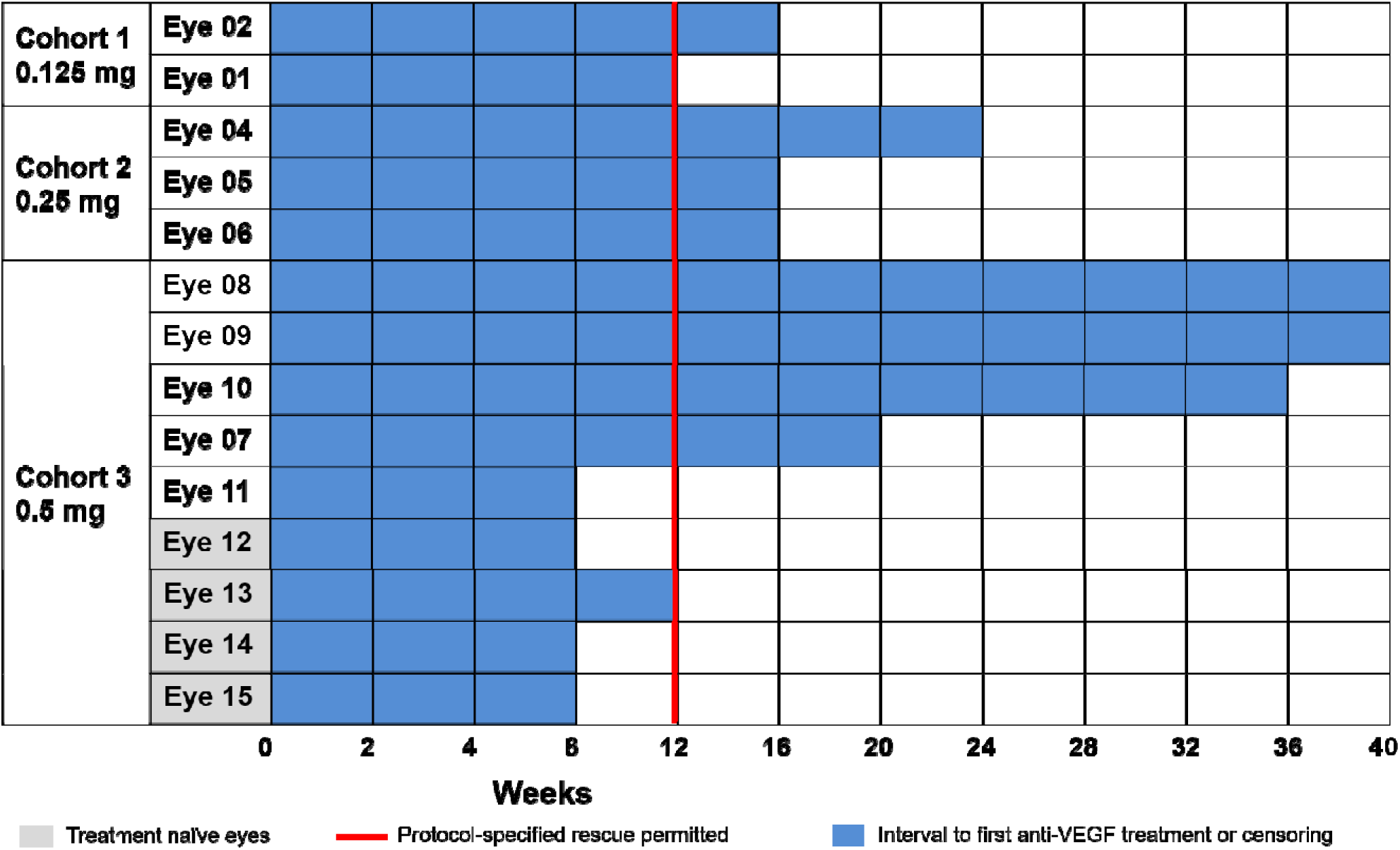
Time to first anti-VEGF treatment or censoring among the 14 eligible eyes. Swimmer plot showing the interval from a single suprachoroidal AXT107 injection to first anti-VEGF treatment or censoring at the last study visit for individual eligible eyes in Cohort 1 (0.125 mg), Cohort 2 (0.25 mg), and Cohort 3 (0.5 mg). Within each cohort, eyes are ordered by time to first anti-VEGF treatment or censoring; treatment-naïve eyes are indicated in gray. Blue bars represent the interval to first anti-VEGF treatment or censoring. The vertical red line marks Week 12, when protocol-specified rescue anti-VEGF treatment became permitted; anti-VEGF treatment administered before Week 12 was investigator initiated.

Among the 9 eyes treated with 0.5 mg, durability appeared greatest in treatment-experienced eyes, 60% of whom remained rescue-free for 8-9 months or through the end of the study at week 40 (median durability of 36 weeks). In contrast, treatment-naïve 0.5-mg eyes generally required earlier protocol-specified rescue and showed more variable clinical courses. In an exploratory analysis among the 14 eligible eyes included in efficacy analyses, longer time since nAMD diagnosis was associated with prolonged durability after AXT107 (Pearson r = 0.52; **Fig. S2**). A second supplemental analysis showed longer treatment-free intervals in treatment-experienced than treatment-naïve eyes, particularly in the 0.5-mg cohort (**Fig. S3**).

### Representative imaging and lesion analysis

OCT images and clinical dashboards representing baseline, the mid-study visit at Week 20, and the last visit before protocol-specified rescue or study completion are shown in **Fig. 5** for treatment-experienced Eyes 08, 09, and 10. These eyes were selected to illustrate prolonged treatment-free intervals or delayed protocol-specified rescue after AXT107. Treatment-experienced Eyes 08 and 09 completed follow-up without protocol-specified rescue, and treatment-experienced Eye 10 remained on AXT107 until Week 36. For each eye, the dashboards display longitudinal BCVA, IOP, and CST measurements over 40 weeks alongside SD-OCT B-scans at baseline, Week 20, and the last study visit before protocol-specified rescue or study completion: Week 40 for treatment-experienced Eyes 08 and 09 and Week 28 for treatment-experienced Eye 10, the last visit at which CST remained lower than baseline before a subsequent increase that preceded rescue at Week 36.

**Fig. 5.**
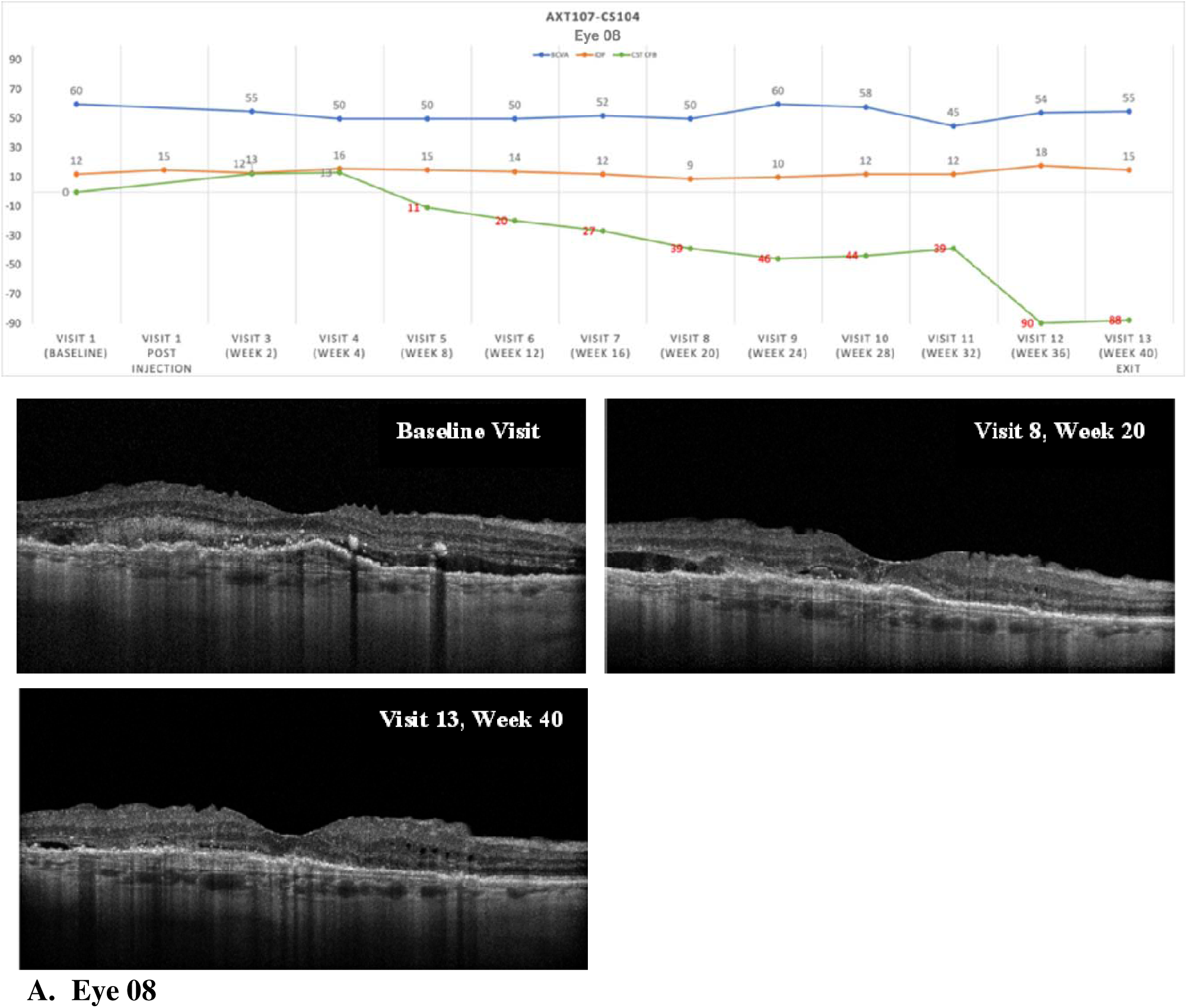
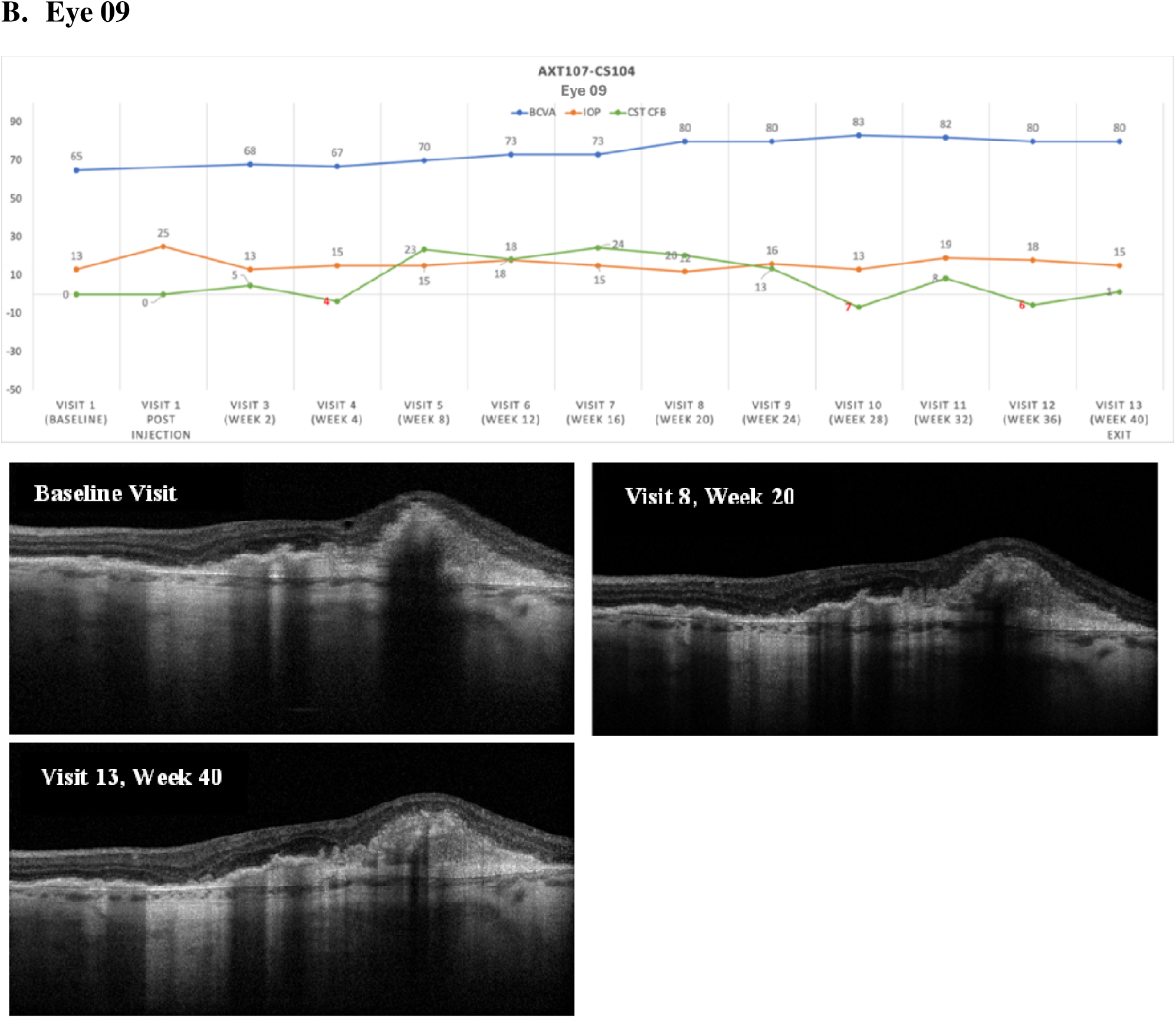
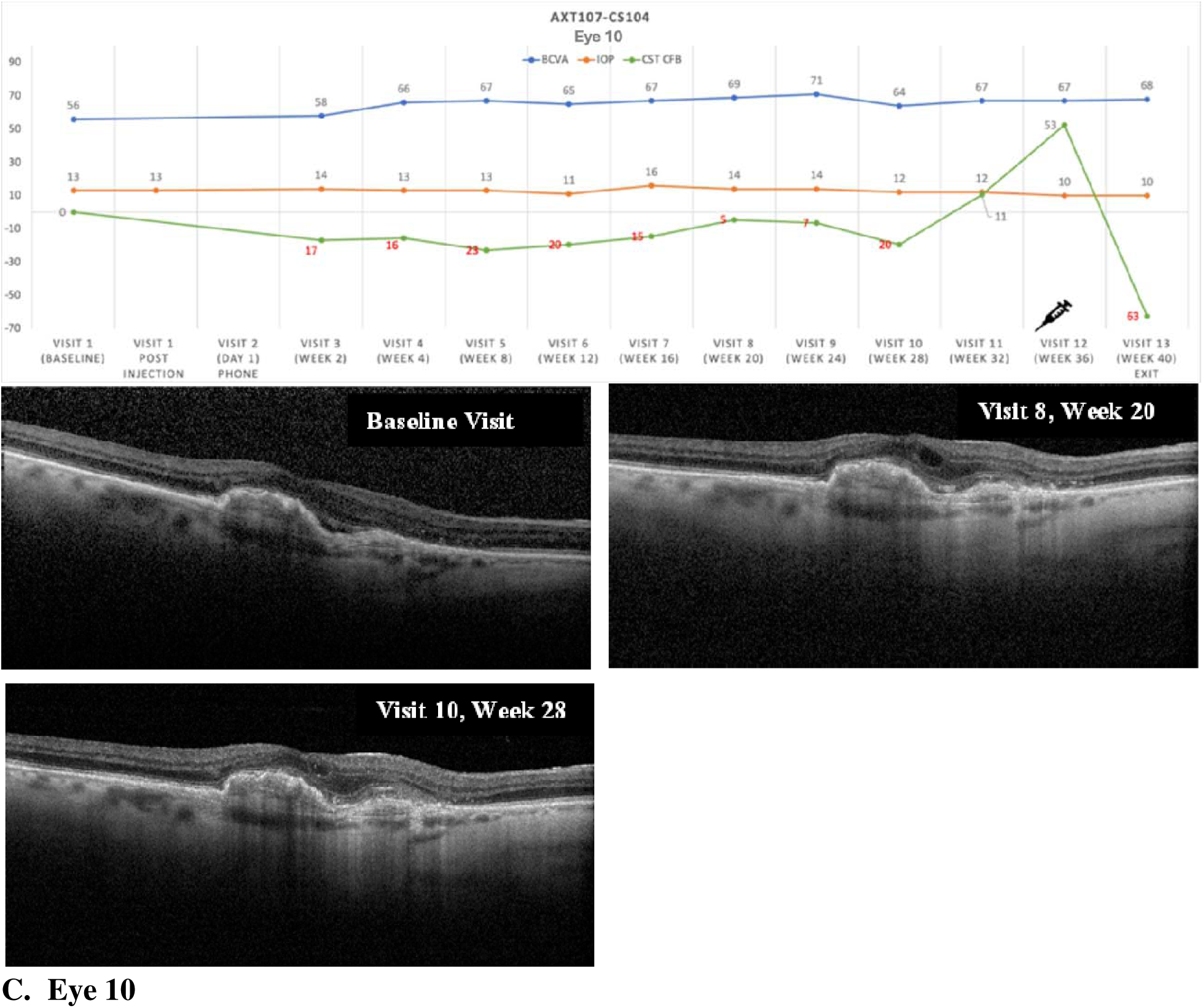
Longitudinal clinical dashboards and SD-OCT imaging in three treatment-experienced Cohort 3 eyes with long treatment-free intervals. (A) Eye 08; (B) Eye 09; and (C) Eye 10. Each panel shows ETDRS letter score, intraocular pressure (IOP), and central subfield thickness (CST) over 40 weeks, with horizontal SD-OCT B-scans at baseline (top left), Week 20 (top right), and the final visit before protocol-specified rescue or study completion (bottom). The final OCT was obtained at Week 40 for Eyes 08 and 09 and at Week 28 for Eye 10, the last visit at which CST remained below baseline before the increase that preceded protocol-specified rescue at Week 36.

## Discussion

DISCOVER was a first-in-human, open-label study of a single suprachoroidal injection of AXT107 microparticles in eyes with nAMD. AXT107 was well tolerated through Week 40, with no increases in intraocular pressure and no cases of endophthalmitis, retinal detachment, suprachoroidal hemorrhage, or clinically significant intraocular inflammation. Two eligible eyes completed Week 40 without protocol-specified rescue anti-VEGF treatment, and one additional eligible eye received protocol-specified rescue at Week 36. To our knowledge, DISCOVER is the first completed clinical study in nAMD to evaluate an integrin-disrupting therapy designed to modulate both VEGFA and angiopoietin-2/Tie2 signaling (18,20), rather than acting through extracellular VEGFA or Ang2 neutralization.

### Safety and route

The most immediate translational result from DISCOVER is that reformulating AXT107 for suprachoroidal delivery appears to have mitigated the safety concern encountered with the earlier intravitreal gel formulation (NCT04746963). No increases in IOP were observed at any dose, and no subject required chronic IOP-lowering therapy. Likewise, no cases of endophthalmitis, retinal detachment, suprachoroidal hemorrhage, or clinically significant intraocular inflammation were seen. Together, these findings are consistent with the rationale that suprachoroidal delivery may reduce exposure of the anterior segment to AXT107 microparticles and thereby mitigate the outflow-related safety concern observed with the earlier intravitreal formulation.

### Durability and activity

A second notable observation was the duration of time before protocol-specified rescue in some eyes after a single administration. Two eligible eyes completed the full 40-week study without protocol-specified rescue, and a third eligible eye first required protocol-specified rescue at Week 36. Changes in CST and BCVA were observed across dose cohorts, but the most favorable combination of durability, CST control, and BCVA maintenance was seen in several eyes treated with 0.5 mg. In an exploratory analysis, mean BCVA change during the AXT107-alone period was more favorable than during the subsequent anti-VEGF treatment period (**Fig. S1**). Because anti-VEGF treatment was initiated in response to clinical course, this observation does not establish comparative efficacy, but the maintenance of vision during the AXT107-alone period supports further evaluation in a controlled trial.

### Treatment-experienced eyes

An exploratory observation that deserves emphasis is that AXT107 appeared particularly promising in treatment-experienced eyes. Within the 0.5-mg cohort, treatment-experienced eyes generally had longer intervals free of protocol specified rescue (median durability of 36 weeks) and more consistent CST and BCVA trajectories than treatment-naïve eyes, although these subgroup comparisons are limited by the very small sample size and non-randomized design. The association between longer-standing disease and later protocol-specified rescue after AXT107 further supports this signal, but it should be interpreted as hypothesis-generating rather than definitive (**Fig. S2**). If confirmed, this pattern would be clinically important because treatment-experienced patients represent a large real-world population with persistent or recurrent exudation despite prior anti-VEGF therapy.

This possible advantage for treatment-experienced patients is biologically plausible given the mechanism of action of AXT107. Unlike VEGF-neutralizing agents alone, AXT107 inhibits VEGF signaling, promotes Tie2 activation in the presence of Ang2, and suppresses vascular inflammation in preclinical models (18,20). Chronically treated nAMD may be especially enriched for the vascular instability, inflammatory tone, and persistent Ang2 signaling that make this multimodal mechanism useful, whereas newly diagnosed eyes may be more variably driven by acute VEGF-dependent exudation. That interpretation remains speculative in this dataset, but it provides a rational framework for future trials enriched for treatment-experienced nAMD patients.

### Limitations and next steps

This study has several important limitations, including its small sample size, open-label design, lack of a control arm, and the availability of investigator-initiated anti-VEGF treatment before Week 12 and protocol-specified rescue anti-VEGF beginning from Week 12 onward, which complicates interpretation of longer-term post-rescue outcomes. The subgroup comparisons between treatment-experienced and treatment-naïve eyes are also limited because only 4 treatment-naïve eyes were enrolled, all at the 0.5-mg dose. It is also important that DISCOVER used relatively stringent washout criteria for prior intravitreal therapy: brolucizumab was not allowed, and prior aflibercept within 8 weeks or ranibizumab or bevacizumab within 6 weeks of baseline in the study eye led to exclusion. Consequently, many treatment-experienced eyes entered the study after a longer interval since their last anti VEGF injection than in some other early phase nAMD trials, which may have contributed to earlier fulfillment of protocol-specified rescue criteria in susceptible eyes and thus to conservative estimates of durability. Even with these design constraints and potential biases toward earlier protocol-specified rescue, the consistency of the safety findings and the durability seen in several individual eyes justify larger controlled studies. Future studies should more rigorously assess long term visual and anatomical outcomes and explore which patient populations, including treatment-experienced nAMD eyes, derive the greatest benefit from AXT107.

## Materials and Methods

### Study Design

DISCOVER (NCT05859776) was a Phase 1/2a, open-label, dose-escalating, multicenter study evaluating the safety, tolerability, bioactivity, and duration of action of a single suprachoroidal injection of AXT107 microparticles in subjects with nAMD. Three sequential dose cohorts were enrolled: Cohort 1 (low dose 0.125 mg/eye), Cohort 2 (mid dose 0.25 mg/eye), and Cohort 3 (high dose 0.5 mg/eye). After an independent Data Monitoring Committee (DMC) reviewed the 14-day safety data from each cohort and issued an ascension vote, the subsequent cohort was opened. Following DMC review and ascension of the first 3 subjects in Cohort 3, this cohort was expanded to include 6 more subjects for a total of 9 subjects. Of the 6 patients enrolled in the Cohort 3 expansion, 4 were treatment-naïve and had newly diagnosed nAMD.

Each subject was followed for 40 weeks after a single injection with scheduled visits at Day 1 (phone), Week 2, Week 4, and every 4 weeks thereafter through Week 40. Study duration, including a 14-day screening phase, was approximately 42 weeks.

### Participants

Eligible subjects were ≥50 years of age with active subfoveal choroidal neovascularization (CNV) of any subtype, or juxtafoveal CNV with leakage affecting the fovea, secondary to AMD with the CNV lesion comprising ≥50% of the total lesion. All subjects were required to have evidence of subretinal or intraretinal fluid on spectral-domain optical coherence tomography (SD-OCT) and a screening BCVA in the study eye between 25 and 65 ETDRS letters (20/320–20/50). Eligibility was determined using the screening assessment. One treatment-naïve study eye, Eye 13, had a screening BCVA of 65 ETDRS letters and was therefore eligible; BCVA was 69 ETDRS letters at the baseline visit approximately 2 weeks later. For Cohort 1, Cohort 2, and the first 3 subjects of Cohort 3, all subjects were required to have been prior partial responders to anti-VEGF therapy. Treatment-naïve subjects were permitted only in the expansion phase of Cohort 3. Key exclusion criteria included non-response to prior anti-VEGF therapy, prior use of brolucizumab, other concurrent ocular diseases requiring surgical or medical intervention, non-AMD causes of CNV, and active uveitis or ocular infection. Additionally, prior aflibercept use within 8 weeks or ranibizumab or bevacizumab within 6 weeks of baseline in the study eye was exclusionary. Post-study review of screening and baseline SD-OCT images identified one treatment-experienced eye, Eye 03, without definite intraretinal or subretinal fluid and therefore not meeting the protocol-specified imaging eligibility criterion. This eye was retained in the safety population because it received AXT107 and completed follow-up but was excluded from all efficacy, durability, rescue, and exploratory analyses. The activity-evaluable population therefore comprised 14 eligible eyes.

### Study drug administration

AXT107 microparticles were prepared as a self-assembled peptide suspension without polymeric excipients by KABS (St-Hubert, Canada) (22) and supplied as a sterile, preservative-free suspension for suprachoroidal injection. Suprachoroidal injections were performed using a precision etched silicon microneedle (Bella Vue, Uneedle, Enschede, Netherlands), which has an ultrashort 0.16 mm bevel designed to localize the needle opening to the suprachoroidal potential space rather than traversing multiple ocular layers. All subjects, except for one mid-dose subject, were injected using a 900 µm Bella-Vue silicon microneedle; one mid-dose subject injected using a 1100 µm needle. Injections were performed by trained retinal specialists at four clinical sites.

### Outcome measures

The primary outcome was safety and tolerability, assessed by the incidence of ocular and systemic adverse events (AEs), serious adverse events (SAEs), and changes from baseline in intraocular pressure (IOP), visual acuity, and ophthalmic examination findings through Week 40. Secondary outcomes included mean change in central subfield thickness (CST) as measured by SD-OCT and mean change in BCVA as measured by ETDRS chart. Additional exploratory analyses included time to first protocol specified rescue anti-VEGF injection and total number of rescue injections through last patient last visit, as well as descriptive proportions of eyes gaining ≥5, ≥10, and ≥15 ETDRS letters at the last on treatment visit (week 40 for eyes without rescue and the rescue day visit for eyes with rescue).

Protocol-specified rescue therapy with the investigator’s preferred intravitreal anti-VEGF agent was permitted starting at Week 12 for any subject meeting pre-specified rescue criteria: ≥9 letter decrease in BCVA from baseline or ≥75 µm increase in CST compared to the lowest measurement at any previous visit.

### Statistical Analysis

All 15 participants who received AXT107 were included in safety analyses. The activity-evaluable population comprised the 14 treated eyes that met the protocol-specified baseline imaging eligibility criterion; Eye 03 was excluded from efficacy, durability, rescue, and exploratory analyses. Safety events and categorical outcomes were summarized as counts and percentages. BCVA and CST outcomes were summarized descriptively using individual-eye measurements and changes from baseline. For analyses of outcomes during the AXT107-treatment period, the assessment immediately before the first subsequent anti-VEGF injection was included, and later assessments were excluded. Time to first anti-VEGF treatment included either investigator-initiated treatment before Week 12 or protocol-specified rescue treatment from Week 12 onward. The exploratory association between time since nAMD diagnosis and time to first anti-VEGF treatment was assessed using Pearson’s correlation and an unadjusted linear regression fit. Given the small, open-label study without a control group, exploratory outcomes were interpreted descriptively rather than as estimates of comparative treatment efficacy.

## List of Supplementary Materials

Fig. S1. Change from baseline BCVA while on AXT107 treatment and during the anti-VEGF rescue period.

Fig. S2. Exploratory association between time since neovascular age-related macular degeneration (nAMD) diagnosis and time to first rescue anti-vascular endothelial growth factor (anti-VEGF) injection.

Fig. S3. Durability of response by dose in treatment-experienced eyes.

## Data Availability

All data produced in the present study are available upon reasonable request to the authors

## Funding

The scientific development of AXT107 was supported by internal funding from AsclepiX Therapeutics and National Institutes of Health grants R01EY028996 and R44EY027632.

## Author Contributions

Methodology: NBP, ACM, TJR

Investigation: DRPA, SD, DRL, WZB

Formal analysis: NBP, ACM, PAC, ASP, JJG

Writing - original draft: NBP

Writing - review & editing: NBP, ACM, TJR, DRPA, SD, DRL, WZB, PAC, ASP, JJG

## Competing Interests

J.J.G. and A.S.P. were co-founders and served as officers and consultants of AsclepiX Therapeutics that conducted the clinical trials described therein; N.B.P. served as VP for R&D at AsclepiX; A.C.M. was a Senior Scientist at AsclepiX; J.J.G., A.S.P. and N.B.P. are co-founders of OptaNova Pharma that acquired AsclepiX’s IP. A.S.P. is a co-founder of Terebra Therapeutics, a consultant to J&J; he receives research funding from Pfizer and Merck, all unrelated to this manuscript. N.B.P. is a co-founder and CEO of Terebra Therapeutics and he is a consultant to Cascade Prodrug. P.A.C.: Personal financial interest in Aerpio, Allegro Ophthalmics, AsclepiX, Ashvattha, Bausch & Lomb, Catawba Research, Celanese, Clearside Biomedical, Cove, CureVac, Exegenesis Bio, Exonate, Genentech, Inc./Roche, Graybug, Gyroscope, Janssen Research & Development, LLC, Merck, Novartis, Perfuse, Wave Life Sciences; Financial support from Aerpio, Ashvattha, Genentech, Inc./Roche, Graybug, Mallinckrodt, Oxford Biomedica, Regeneron, Regenxbio, Sanofi Genzyme. The terms of these arrangements are being managed by the Johns Hopkins University in accordance with its conflict-of-interest policies. D.R.P.A: D.R.P.A has received research funding or free research materials (including equipment) or services from the following companies: AbbVie, Acelyrin, Aerie, Alcon, Aldeyra, Alimera Sciences, Allergan, Alumis, Annexon, Apellis, Ascidian, AsclepiX, Ashvattha Therapeutics, Aura, Aviceda Therapeutics, Bayer, Beyeonics, Boehringer Ingelheim, Breye, Character Biosciences, Cognition Therapeutics, Dutch Ophthalmics, Eluminex Biosciences, Exegenesis Biosciences, EyeBio, Eyepoint Pharmaceuticals, Galimedix, Genentech, Gyroscope, i-Lumen Scientific, Invirsa, Iveric Bio, Janssen, Kodiak Biosciences, Novartis, Ocular Therapeutix, Ocugen, Oculis, ONL Therapeutics, Opthea, Palatin,, Priovant, RegenexBio, Regeneron, Roche, Samsara Vision, Sana Holdings, Sanofi, SmileBiotek, Unity Biotechnology, and Zeiss. S.D.: S.D. has received funding from Genentech, Regeneron, Bausch & Lomb, Harrow, Amgen, and 4Front. D.R.L.: D.R.L. D.R.L. has received research funding from 4DMT, Affamed Therapeutics, Alexion, Annexon Biosciences, AsclepiX Therapeutics, Astellas, Aviceda, Biogen, Eyebio, Eyepoint Pharmaceuticals, Genentech, Kalaris Therapeutics, Kodiak Biosciences, Notal Vision, Ocular Therapeutix, ONL Therapeutics, Opthea, Opus Genetics, and Regeneron; he is a consultant to 4DMT, Annexon Biosciences, Biogen, Ashvattha Therapeutics, Astellas, Boehringer Ingelheim, Eyepoint Pharmaceuticals, Kriya Therapeutics, Thea Pharma, Neurotech, Ocular Therapeutix, Opthea, Opus Genetics, Regeneron, and Stealth Biosciences; he has held stock/equity options with Eyepoint Pharmaceuticals, Kriya Therapeutics, and Ocular Therapeutix. W.Z.B.: W.Z.B is a consultant for 4DMT. TJR has no competing interests to declare.

## Data and Materials Availability

Quantitative datasets underlying the figures in this report, including longitudinal raw BCVA values, change-from-baseline BCVA trajectories, detailed CST measurements, and the SD-OCT images underlying the reported analyses and representative images, will be deposited in a dedicated Johns Hopkins University database and assigned a persistent DOI. The DOI and access instructions will be provided to journal reviewers during peer review and made available to the broader community upon publication, enabling interested readers to access and re analyze the full dataset.

## Notes

### Clinical Trial

NCT05859776

### Author Declarations

Advarra IRB gave ethical approval for this work. Compliance Statement/REB Attestation (Applicable for research conducted in Canada): The IRB attests that this submission has been approved by an IRB whose membership complies with the requirements defined in Health Canada regulations, ICH GCP guidelines, FDA regulations at 21 CFR part 56, and HHS regulations at 45 CFR part 46. The IRB carries out its functions in accordance with FDA regulations at 21 CFR parts 50, 56, 312, and 812; HHS regulations at 45 CFR part 46, subparts A-E; good clinical practices; Health Canada regulations; and the Tri-Council Policy Statement: Ethical Conduct for Research Involving Humans, as appropriate to the research. (For NHP studies only) Please note that this research has been reviewed by a person knowledgeable in complementary and alternative health care, who <participated at the IRB meeting and/or submitted written comments>;. Advarra IRB is registered with OHRP and FDA under IRB#00000971.

### Summary of Updates

Author Thomas J. Robinson was inadvertently left out in my earlier submisson. I have now added his name, affiliation and ORCID ID.

