## Supplemental data for "Single-Dose Suprachoroidal AXT107 in Neovascular Age-Related Macular Degeneration: The Phase 1/2a DISCOVER Trial" (from another version of this preprint: version 1, 2026-09-24)

### Supplementary Figures

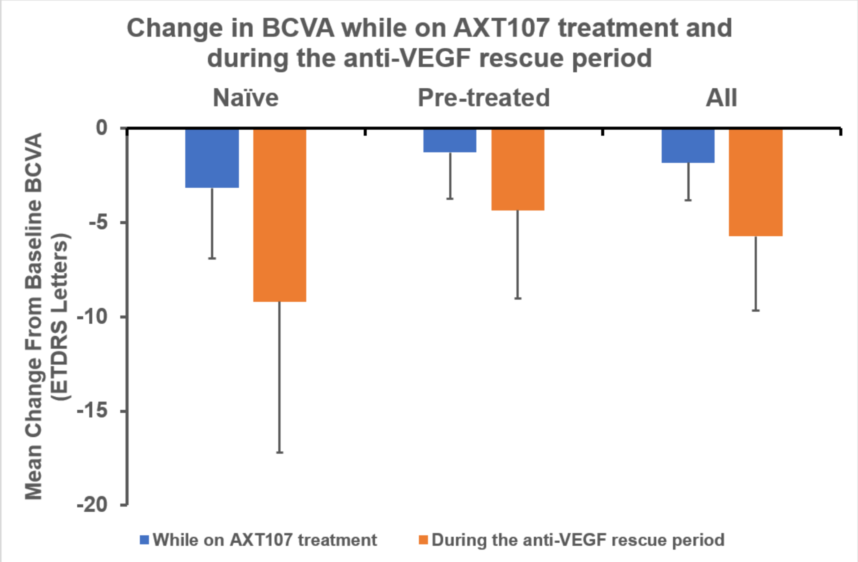


**Supplementary Figure 1. Change from baseline BCVA while on AXT107 treatment and during the anti-VEGF rescue period.**

Mean change from baseline best-corrected visual acuity (BCVA), in ETDRS letters, is shown for treatment-naïve eyes, treatment-experienced eyes, and all eligible eyes. For eyes receiving rescue anti-VEGF treatment, the BCVA assessment obtained at the first rescue visit before injection was included in the AXT107-treatment period; the anti-VEGF rescue period included BCVA assessments at subsequent visits through Week 40 or the last available visit. For eyes not receiving rescue anti-VEGF treatment, BCVA assessments through Week 40 or the last available visit were included in the AXT107-treatment period. The AXT107-treatment period included 4 treatment-naïve eyes, 10 treatment-experienced eyes, and 14 eligible eyes overall; the anti-VEGF rescue-period analysis included 4, 8, and 12 eyes, respectively. Bars represent means, and error bars represent the standard error of the mean (SEM). All efficacy analyses excluded treatment-experienced Eye 03, which did not meet the protocol-specified baseline imaging eligibility criterion. This exploratory descriptive analysis was not designed to compare treatment effects because rescue anti-VEGF treatment was initiated according to clinical course.

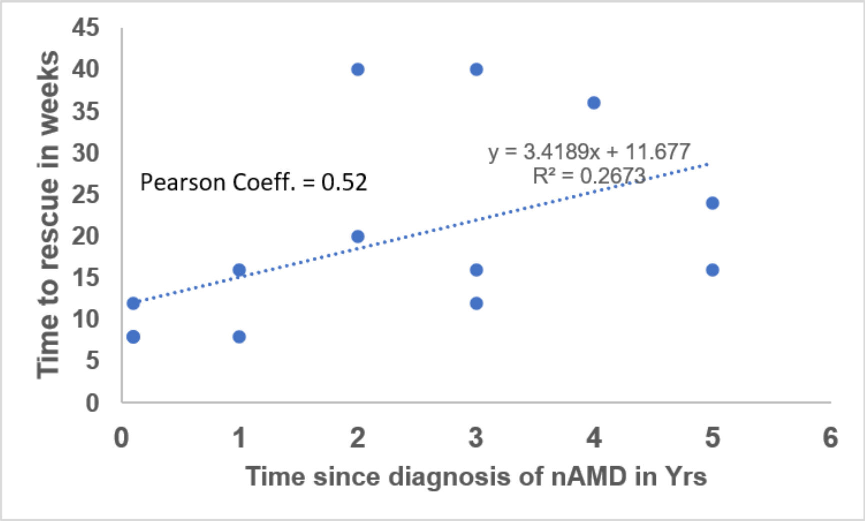


**Supplementary Figure 2. Exploratory association between time since neovascular age-related macular degeneration (nAMD) diagnosis and time to first rescue anti-vascular endothelial growth factor (anti-VEGF) injection.**

Each dot represents one eligible eye (n=14). Four treatment-naïve eyes were newly diagnosed with nAMD and had a recorded time since diagnosis of 0 months. Of these, three eyes had the same time to first rescue (8 weeks) and are superimposed at the same plotted coordinates. The dotted line indicates the unadjusted linear regression fit. Eye 03 was excluded because it did not meet the protocol-specified baseline imaging eligibility criterion. This modest association should be interpreted as hypothesis-generating.

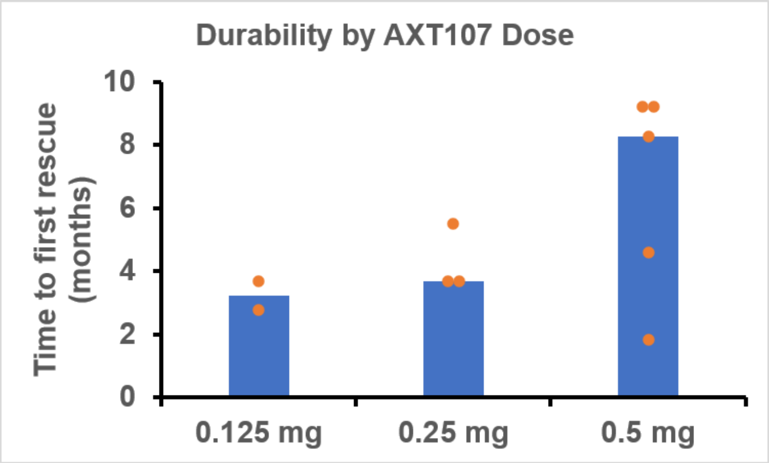


**Supplementary Figure 3. Durability of response by dose in treatment-experienced eyes.**

Time to first protocol-specified rescue anti-VEGF injection, or censoring at Week 40 for eyes without rescue, is shown by AXT107 dose cohort. Blue bars indicate the median treatment-free interval, and orange circles indicate individual eyes. The treatment-experienced 0.5 mg cohort had the longest treatment-free interval, with a median of 36 weeks (approximately 8.3 months); two eyes remained rescue-free for 8-9 months or through Week 40.
